# Towards Inpatient Discharge Summary Automation via Large Language Models: A Multidimensional Evaluation with a HIPAA-Compliant Instance of GPT-4o and Clinical Expert Assessment

**DOI:** 10.1101/2025.04.03.25325204

**Authors:** Tyler Osborne, Sadia Abbasi, Stephanie Hong, Robert Sexton, Jonathan Ambut, Neil J. Patel, Richard N. Rosenthal, Lyncean Ung, Fusheng Wang, Rachel Wong

## Abstract

Large language models (LLMs) have demonstrated potential to automate clinical documentation tasks that may reduce clinician burden, such as generation of hospital discharge summaries. Prior research used older LLMs and limited data, raising concerns about fabrications and omissions. In this study, we evaluated the automatic generation of inpatient Internal Medicine discharge summaries using a HIPAA-compliant Microsoft Azure instance of OpenAI’s GPT-4o. Both human-written and AI-generated discharge summaries were scored by Internal Medicine hospital faculty for quality, readability/conciseness, factuality and completeness, presence of hallucinations/omissions and their impact on safety, and compared with the actual discharge summaries. Our results showed that the AI-generated discharge summaries significantly outperformed actual human written summaries in both quality and readability/conciseness and were comparable to humans in factuality and completeness, with a minimal cost.

## Introduction

Patients are highly vulnerable to experiencing adverse outcomes during the transition of care from the hospital to community settings^1,2^, which can be exacerbated by poor communication between inpatient and outpatient providers at hospital discharge^3^. The discharge summary, which is the primary method for documenting patients’ diagnoses, test findings, hospital courses, and follow-up needs, is a complex and burdensome clinical documentation task for providers that is prone to errors. Studies have found medication discrepancies in 38-94% of discharge summaries for elderly patients^4–6^, high rates of missing^7^ or irrelevant information^8^, and availability rates of discharge summaries as low as 12-34% on follow up^7,9^. Issues with quality and timeliness of discharge summaries can lead to poor patient outcomes, such as adverse medication events, missed appointments or tests on follow up, and hospital readmission^3,7,10,11^. Additionally, clinical documentation in the electronic health record (EHR) is a significant burden for healthcare providers and contributes to physician burnout^12,13^. The Joint Commission requires that hospital discharge summaries be completed within 30 days, with the following components: 1) reason for hospitalization, 2) significant findings, 3) procedures/treatments provided, 4) patient’s discharge condition, 5) patient/family instructions and 6) attending physician’s signature^14^. Poor quality or delay in discharge summary creation can also result in compliance or liability issues for healthcare organizations^14–16^. Although there have been quality improvement efforts to address challenges in creating timely, accurate discharge summaries, they are often human-time intensive, such as resident training, use of multidisciplinary teams, or addition of templates with new sections or writing tips^17–19^. Discharge summary generation is a time consuming, manual task that is prone to errors, and application of technologies such as LLMs are promising avenues to reduce clinician burden and improve the quality of documentation.

### Related Work

There has been recent work in the use of natural language processing and LLMs for automating generation of discharge summaries. In this section, we enumerate some of these recent papers, their methods, results, and takeaways that informed this study. Hartman et al. used a BERT and BART-based approach, relying on gold standard discharge summary text to select via maximized ROUGE scores the most relevant notes to summarize^20^. They found that 62% of the automated summaries met standards of clinical validity, establishing transformer models as effective summarizers of clinical text. Their manual evaluation of four categories (quality, readability, factuality and completeness) identified issues with LLM factuality and completeness, and they also reported errors of fabrication or omission as part of their evaluation framework.

Zaretsky et al. used GPT-4 via Microsoft Azure (HIPAA compliant) to modify 50 human-written discharge summaries to make the language more readable and patient friendly, measuring readability via the Flesch-Kincaid Reading Ease score^21^. Clinical reviewers found that the transformed discharge summaries were significantly more readable and understandable than the original summaries but identified several safety concerns due to omissions and fabrications. While 54% of their AI-generated summaries scored at the highest level of accuracy, 39% were noted to contain potential safety risks. Their work established the feasibility of HIPAA-compliant usage of current-generation LLMs only accessible via API, and reinforced known limitations of LLMs on this task regarding accuracy, completeness and errors.

Schwieger et al. also used GPT-4 to generate 30 standardized discharge summaries drawn from pseudonymized Psychiatry encounters, with blinded evaluation of AI and human-written reports^22^. The manual reviews consisted of Likert ratings on 15 dimensions, the most salient being “As a senior doctor, I would only have to make few corrections,” and “I would send out this discharge summary in its current form.” Using the latter dimension as a proxy for overall quality, the authors found that the human-written summaries were rated significantly higher than the AI-generated ones, surpassing AI’s performance in 12 of 15 dimensions, and 45% of the AI summaries were rated as unfavorable on the “low expected correction effort” dimension. The authors found clinically relevant hallucinations in 40% of the AI-generated summaries, of which 38% were “highly relevant,” and no hallucinations in the human-written summaries. This study provided an important counterargument to the assertion that LLMs have nearly closed the gap between human-written and AI-generated discharge summaries.

Williams et. al. used both GPT-4 and GPT-3.5-turbo to generate discharge summaries from deidentified emergency department examination notes for 100 emergency medicine encounters, performing manual evaluation by clinicians^23^. They found that GPT-4 produced fewer errors than GPT-3.5-turbo, with GPT-4 producing inaccuracies, hallucinations and omissions in 10%, 42% and 47% of the generated summaries, respectively. 47% of the GPT-4 summaries were entirely error-free after excluding certain trivial errors resulting from limitations of the evaluation method. This work showed how employing the latest models can significantly improve performance without any additional methodological adjustments.

Overall, while studies have shown promise for the use of LLMs in generating discharge summaries, limitations to implementation include pragmatic concerns regarding patient privacy and cost, lack of studies using full hospitalization records from real world data, and lack of multi-dimensional evaluation for both quality and safety evaluation. In this project, we used complete sets of real-world electronic health record clinical notes from Internal Medicine (IM) hospital encounters to develop a modern LLM-based model for generating discharge summaries. We addressed several major factors that will inform future implementation, including HIPAA-compliant state-of-the-art LLM services, data cleaning and normalization, evaluation of quality and safety by clinical by subject matter experts and cost considerations.

### Dataset

We extracted all clinical notes, test reports, structured data elements, and discharge summaries from 200 Internal Medicine inpatient hospital encounters ranging from 41 to 120 hours in length of stay (average 80, SD ± 20) between January 1 to December 31, 2023, at Stony Brook University Hospital (SBUH), a tertiary care academic center in Long Island, New York. We excluded any encounters with total chart character counts of greater than 120,000 to accommodate GPT-4o’s 128000-token context window length. While one character does not necessarily equal one token, we used this heuristic as an upper bound to reasonably limit the amount of review required of the clinicians performing manual evaluations. A total of 108 encounters were included in our study, with patient representation of 52% (56/108) female and 48% (52/108) male. Patient age varied from 25 to 98 years, with an average age of 67, SD ± 19.

For each encounter, we extracted all the related clinical notes from the hospital’s EHR database, of which there were 545 unique types in the following major categories: ED notes, Consult Notes, Admission and History & Physical Notes, Progress Notes, Addendums and Event Notes. Based on a review of note types by a practicing physician informaticist for clinical relevance, 20% (111/545) of the total note types were included as inputs. The average number of note types used per encounter was 18, SD ± 5. Figure 1 summarizes our dataset creation pipeline.

**Figure 1.**
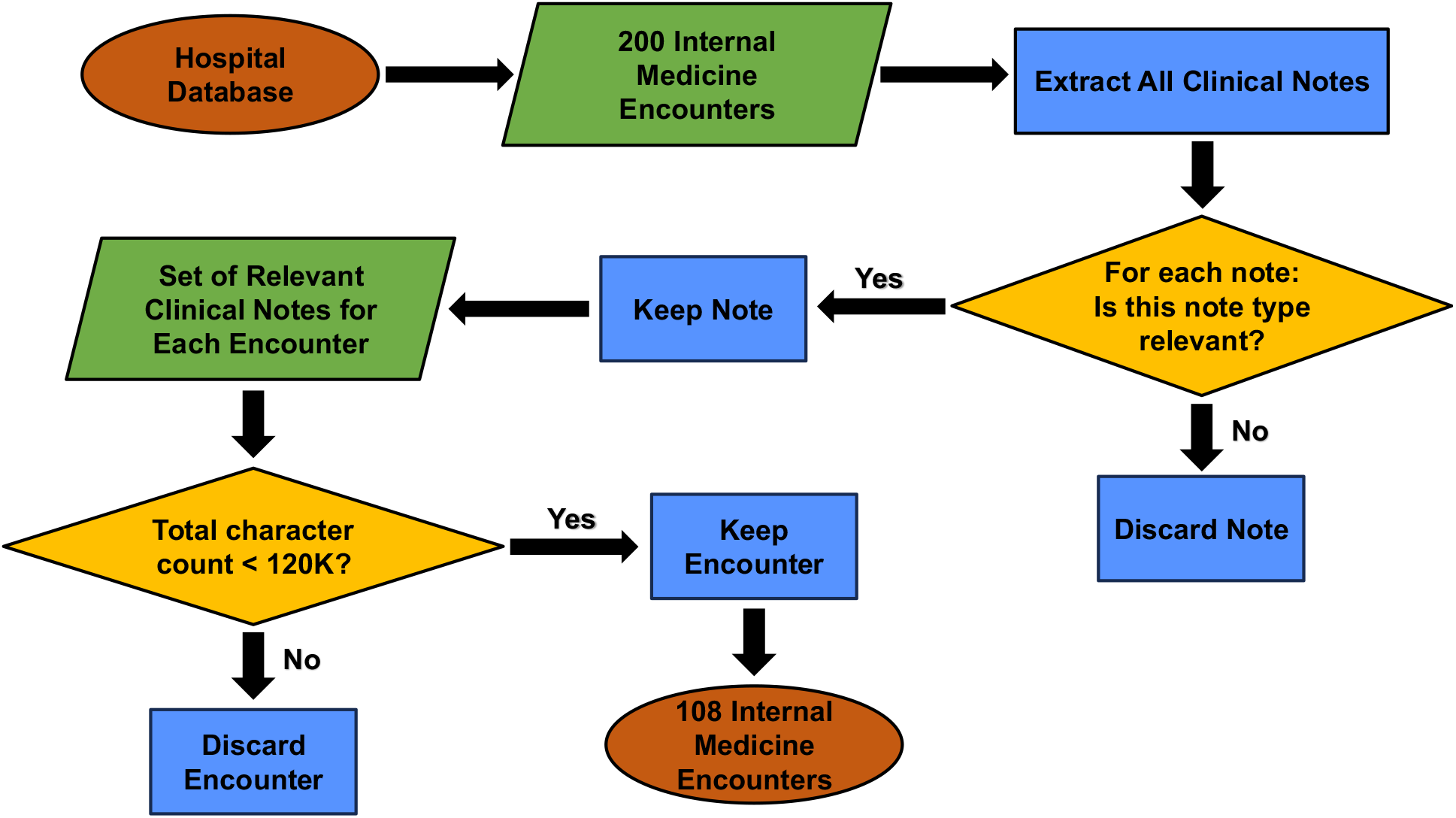
Pipeline diagram illustrating our clinical note extraction process, yielding 108 IM encounters and their relevant clinical notes for use in this study.

Once processed, we used SQLite, a lightweight SQL database engine to store the 108 IM encounters and their associated relevant clinical notes to enable versatile data manipulation via SQL queries.

## Methods

### HIPAA-Compliant Environment to Protect Private Health Information

To maintain HIPAA compliance, all computation was performed via secure shell (SSH) on an in-network computing cluster at SBUH; the data were housed on the same cluster. All inputs to GPT-4o were submitted via API to a private HIPAA-compliant Microsoft Azure OpenAI Service instance maintained by SBUH’s Department of Biomedical Informatics and verified to be safe for processing private health information (PHI) by the Stony Brook Medicine Information Technology team. Codebase version control was performed using a private GitHub instance, also hosted inside the hospital network. Manual clinician evaluations were performed within a secure virtual desktop interface and recorded in a Microsoft Form created inside the SBUH firewall. The study was approved as human subjects research by the Stony Brook University Office of Research Compliance.

### Data Preprocessing

The notes extracted from Oracle’s EHR system were in the RTF format, with the header containing PHI data such as patient names and medical record numbers. To minimize the use of PHI in our study, we implemented a structured pre-processing pipeline for extracting plaintext and de-identifying clinical notes. The pipeline removed all direct identifiers but did not guarantee the removal of indirect identifiers; processed notes were still treated as identifiable data. 1) Plaintext Extraction and Standardization. To standardize the text for LLM processing, we used the `striprtf` Python library to extract plaintext and convert all text to lowercase. This ensured consistency in text representation for LLM processing. 2) Name Extraction and De-identification. Regular expression-based methods were insufficient to extract personal names, so we utilized an LLM to accurately identify all patient and provider names within the notes and used string matching to remove all instances of names.

### Parallel Processing for Efficiency

We used Python 3.12 to perform the computation in this study. To improve efficiency, we implemented parallel processing with Python’s `multiprocessing` library (`starmap` function) as well as the `asyncio` library for sending asynchronous API requests to Microsoft Azure. This boosted the performance by a time factor of 6 with 12 CPU cores.

### Experimental Setup

To produce the AI-generated discharge summaries, we used zero-shot prompting, inserting the clinical notes as context to the model. Following a system prompt outlining the discharge summary generation task, we sorted all the relevant notes for each encounter in temporal order according to their timestamps. These notes were then combined into a single string, with custom headers at the top of each note indicating the note type, its timestamp and clear delimiters between the start and end of each note’s body text. Figure 2 contains the system prompt we used to generate the discharge summaries, and figure 3 illustrates our full preprocessing and output generation pipeline.

**Figure 2.**
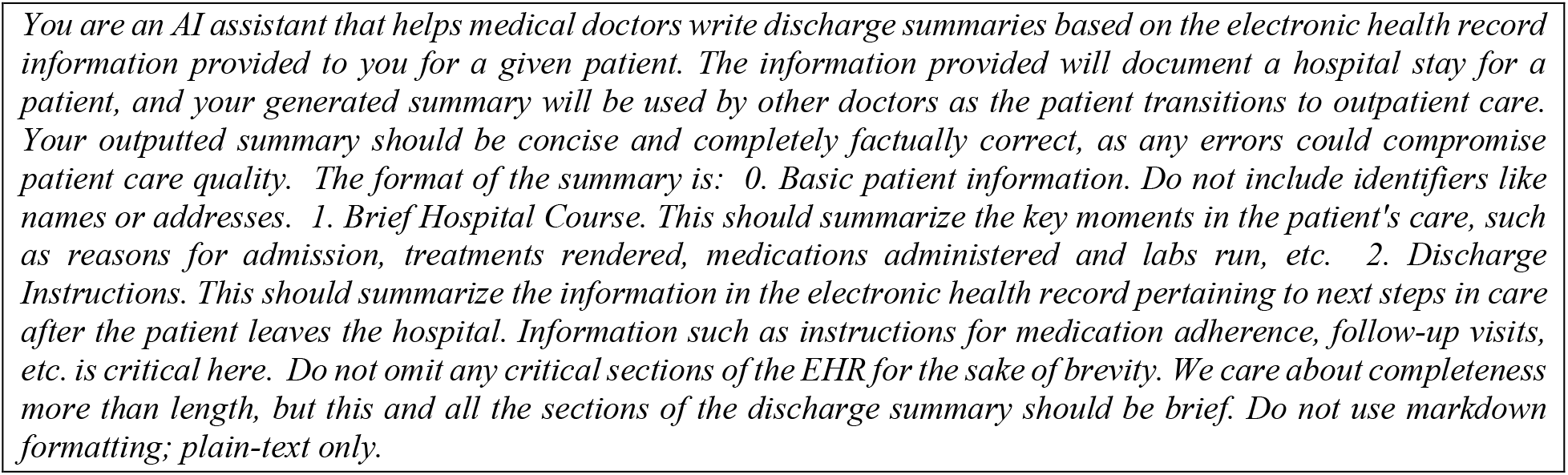
System prompt given to GPT-4o for generating the AI discharge summaries in this study.

**Figure 3.**
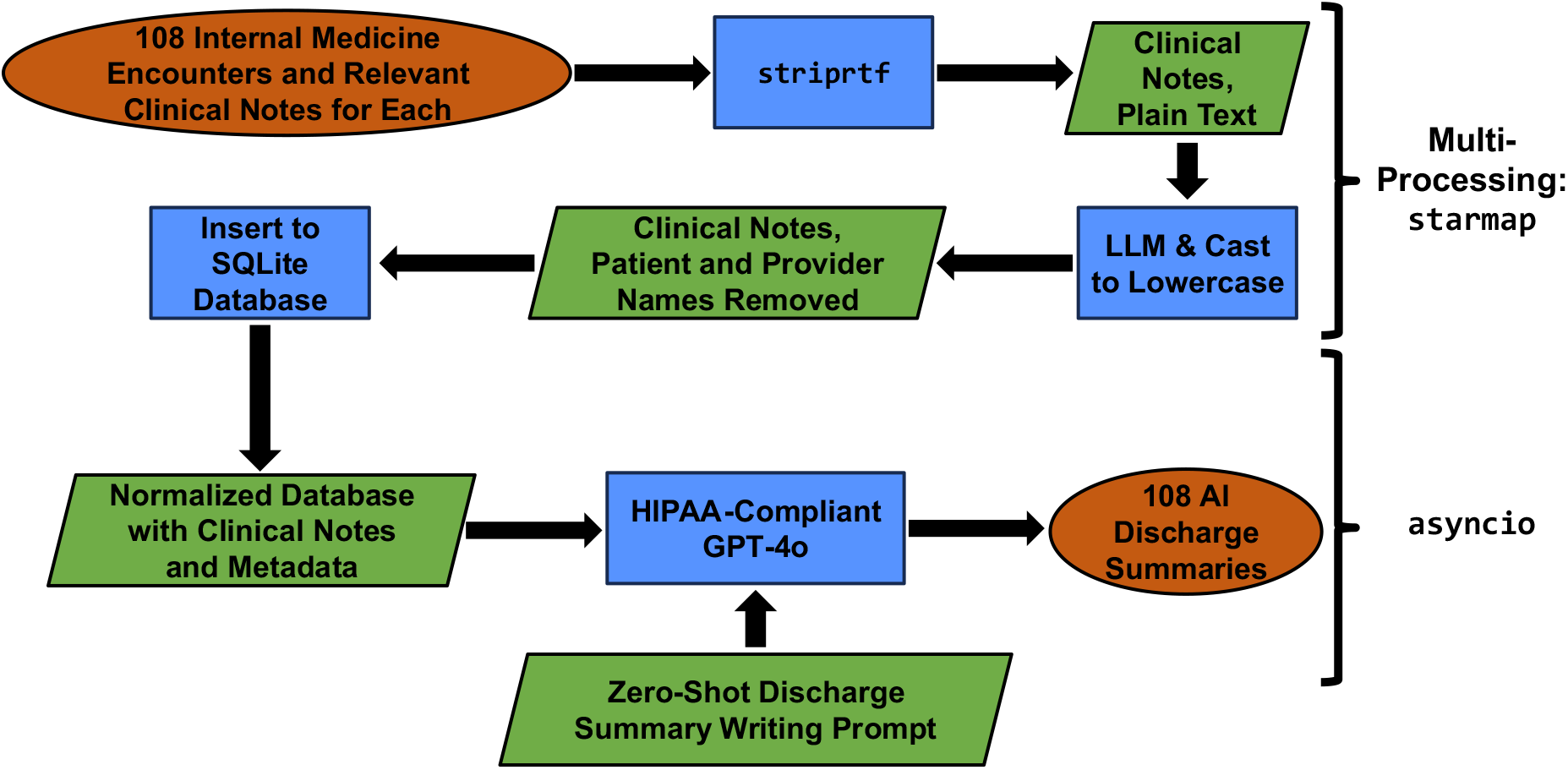
Pipeline diagram for data preprocessing and AI discharge summary generation.

Figure 4, below and continuing onto the next page, shows an example of an AI-generated discharge summary based on the clinical notes written by a clinician for a fictional encounter.

**Figure 4.**
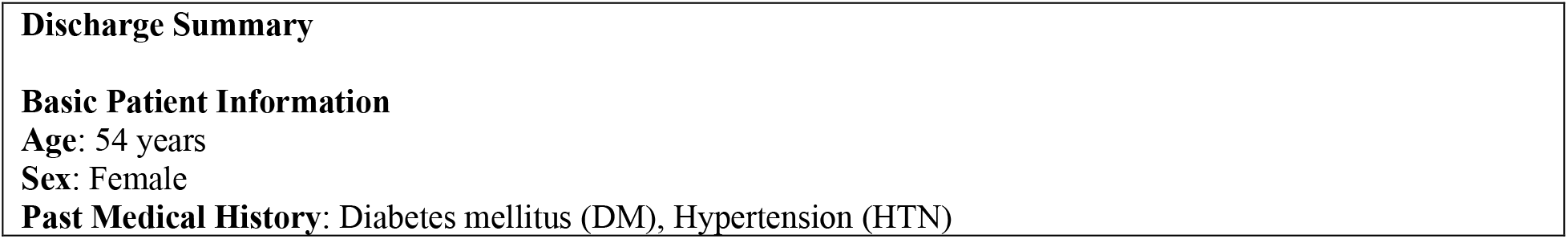
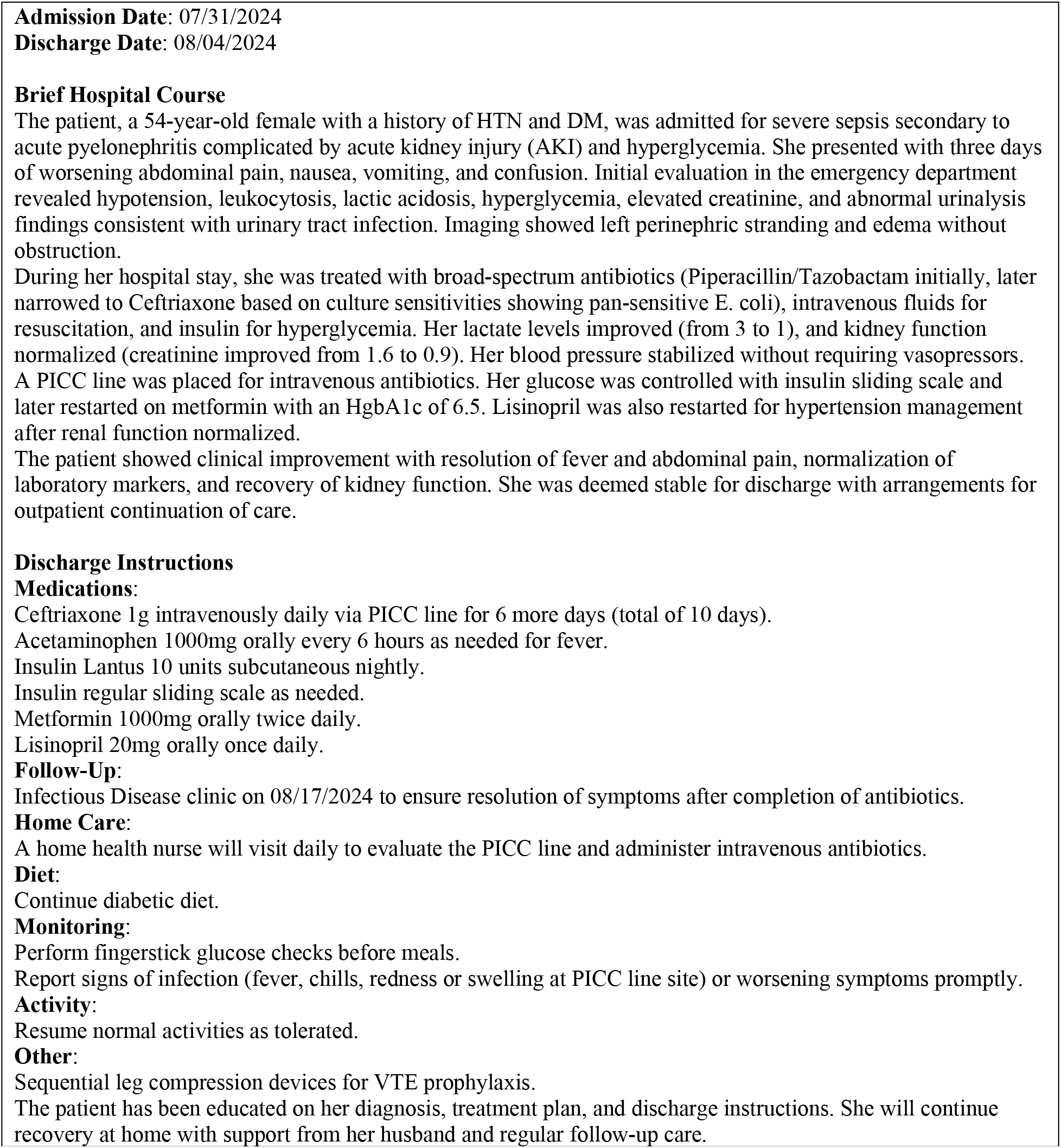
Example of an AI-generated discharge summary based on the clinical notes written by a clinician for a fictional encounter.

### Manual Evaluation

Four practicing Internal Medicine hospitalists evaluated 28 of the 108 AI-generated and human-written discharge summaries, with 7 encounters assigned to each clinician. Each of the 28 encounters was evaluated once. Evaluation consisted of two parts. First, clinicians provided Likert scale ratings, from 1 (low) to 7 (high) with 4 being neutral, on four dimensions: quality, readability/conciseness, factuality and completeness^20^. Ratings were collected for both the human-written and AI-generated discharge summaries. Table 2 describes each rating category.

**Table 2.** Descriptions of Likert rating dimensions for manual clinical expert evaluations.

| Likert Rating Dimension | Description |
| --- | --- |
| Quality | Rate the summary of what you view as a well-written discharge summary. Keep in mind that the intended purpose of the discharge summary, as set out by CMS and the Joint Commission, is to clearly communicate a patient's care plan to the post-hospital care team. |
| Readability/<br>Conciseness | The language and grammar are what would be expected of a trained physician. The intended audience for the summary is the post-hospital care team (usage of medical terminology, abbreviations, and syntax is acceptable if a general physician could understand them). |
| Factuality | All information presented is accurate and factually correct. |
| Completeness | The summary includes the relevant clinical events that occurred during the patient stay. Assess the summary on the standard for not missing any key clinical information that would be detrimental to the patient's post hospital care if the downstream provider were not informed. |

The safety evaluation asked the clinicians to identify instances of fabrication (i.e., incorrect information) and omission (i.e., missing information) that could result in patient harm. This also consisted of four dimensions: a brief description of the issue or error, the error type, the likelihood of patient harm, and the extent of patient harm^24^. Table 3 enumerates the possible values for each dimension.

**Table 3.** Possible values for recording instances of fabrication and omission for manual clinical expert evaluations.

| Dimension | Possible Values |
| --- | --- |
| Issue/Error | Free Text |
| Error Category | Fabrication, Omission |
| Likelihood of Patient Harm | Low, Medium, High |
| Extent of Patient Harm | None, Mild/Moderate, Severe/Death |

For example, an error might be recorded as “former smoker; fabrication; low; none” or “forgot antibiotics on discharge; omission; medium; severe/death.”

## Results

### Manual Evaluation: Likert Ratings and Significance Testing

Table 4 presents summary statistics of the Likert rating data from the clinical expert ratings of 28 human-written and AI-generated discharge summaries, respectively.

**Table 4.**
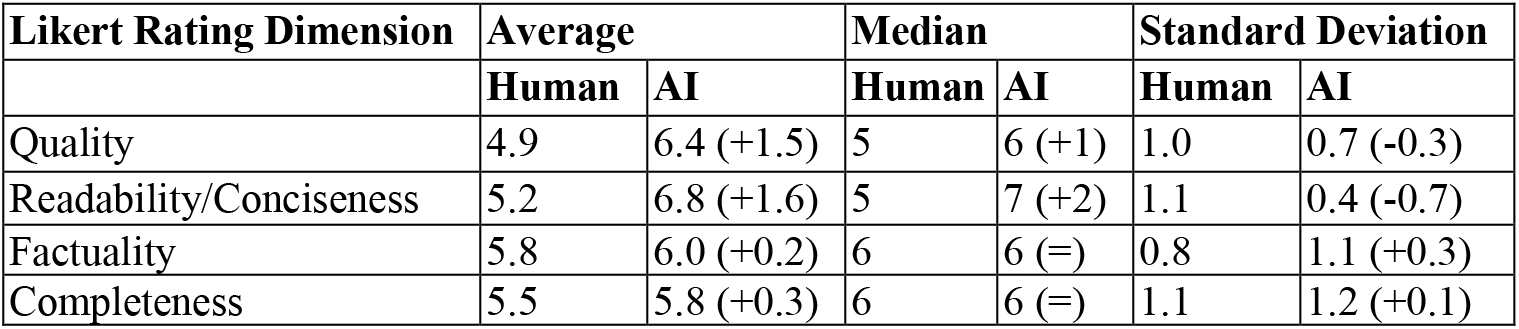
Average, median and standard deviation values for clinical expert ratings of 28 human-written and AI-generated discharge summaries. Likert scale was 1 (low) to 7 (high) with 4 being neutral. Note that between the Human and AI subcategories for each statistic, the values are not paired with respect to individual patient encounters; they are global. The values in parentheses for the AI subcategory represent the delta from the value in the corresponding human subcategory.

We conducted significance testing using a one-sided Wilcoxon paired sample test. The null hypothesis was that the distributions of Likert ratings would be the same for both the human and AI-generated summaries, and the alternative hypothesis was that the distributions of Likert ratings for the human-written summaries, per category, would be less than those of the AI-generated ones. For quality and readability/conciseness, the null hypothesis was rejected with *p* ≪ 0.01, while for factuality and completeness, it was not rejected, with *p* = 0.19 for both categories. With ∝ = 0.05, the AI-generated summaries were rated significantly better than the human-written ones on the first two categories, and for the latter two categories, they were rated similarly.

### Manual Evaluation: Fabrication/Omission Detection

Tables 5 and 6 present respective counts of fabrications and omissions, separated by likelihood and extent of patient harm, for the clinical expert evaluations of 28 human-written and AI-generated discharge summaries.

**Table 5.** Counts of fabrications for 28 human-written and AI-generated discharge summaries as identified by clinical experts. Note that between the Human and AI subcategories, the values are not paired with respect to individual patient encounters; they are global.

|  | Extent of Harm | None |  | Mild/Moderate |  | Severe/Death |  |
| --- | --- | --- | --- | --- | --- | --- | --- |
|  |  | Human | AI | Human | AI | Human | AI |
| Likelihood of Harm | Low | 1 | 5 | 1 | 1 | 0 | 0 |
|  | Medium | 0 | 0 | 2 | 3 | 0 | 0 |
|  | High | 0 | 0 | 0 | 0 | 0 | 0 |

**Table 6.**
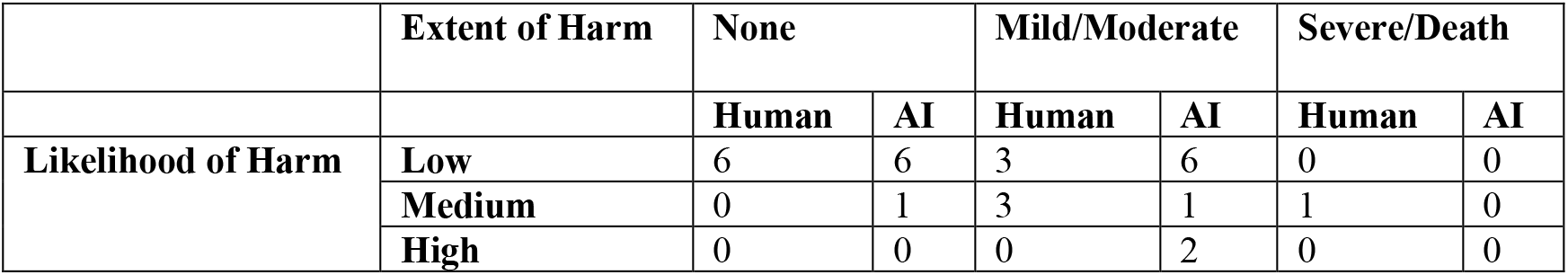
Counts of omissions for 28 human-written and AI-generated discharge summaries as identified by clinical experts. Note that between the Human and AI subcategories, the values are not paired with respect to individual patient encounters; they are global.

We found that neither human-written nor AI-generated summaries contained any fabrications with a high likelihood of severe patient harm, but that AI-generated summaries had about twice as many total fabrications. Importantly, most of the AI fabrications were rated as having a low likelihood of occurrence and no associated patient harm; within the mild/moderate extent of harm category, the counts were similar between the human-written and AI-generated summaries.

We observed similar overall performance between the human-written and AI-generated summaries in terms of the total number of omissions (13 vs. 16), and no AI-generated omissions resulting in severe patient harm.

Table 7 presents the same counts of fabrications and omissions, categorized by the kind of error: medication, diagnosis, test result, and follow up/recommendations.

**Table 7.** Counts of fabrications and omissions categorized by the kind of error. Note that between the Human and AI subcategories, the values are not paired with respect to individual patient encounters; they are global.

| Error Category | Medications |  | Diagnoses |  | Test Results |  | Follow-Up/<br>Recommendations |  |
| --- | --- | --- | --- | --- | --- | --- | --- | --- |
|  | Human | AI | Human | AI | Human | AI | Human | AI |
| Count of Fabrications | 2 | 2 | 0 | 1 | 0 | 2 | 2 | 4 |
| Count of Omissions | 1 | 4 | 5 | 5 | 2 | 2 | 5 | 5 |

For both the human-written and AI-generated discharge summaries, we found that the most common error category identified by the clinician reviewers was related to follow-up/recommendations, followed in descending order by diagnoses, medications and test results.

### Cost Analysis

In this section, we present summary statistics for the cost, in US dollars, to generate 108 AI discharge summaries via the Azure API. As of writing this paper, for GPT-4o, the on-demand Azure API cost per one million un-cached input tokens is $2.50 ($0.0000025 per token or $1 per 400,000 tokens), and $10 per one million output tokens ($0.00001 per token or $1 per 100,000 tokens); we consider all inputs to be un-cached. We used the `tiktoken` library in Python to calculate the token counts. Table 8 presents summary statistics for the number of input and output tokens for 108 AI-generated discharge summaries and the associated API costs.

**Table 8.**
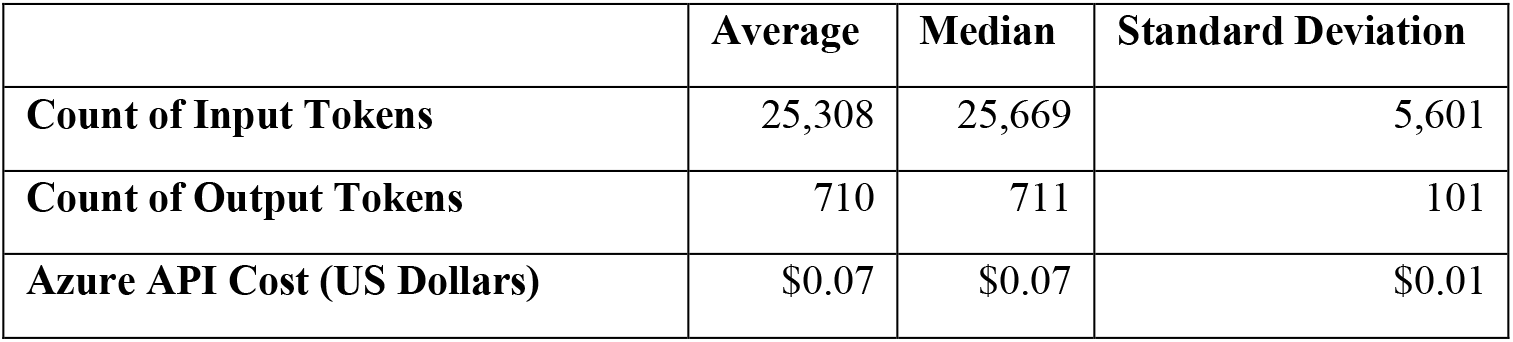
Average, median, and standard deviation values for the counts of input and output tokens for 108 AI-generated discharge summaries and the associated API costs. The cost values combine input and output costs.

|  | Average | Median | Standard Deviation |
| --- | --- | --- | --- |
| Count of Input Tokens | 25,308 | 25,669 | 5,601 |
| Count of Output Tokens | 710 | 711 | 101 |
| Azure API Cost (US Dollars) | \$0.07 | \$0.07 | \$0.01 |

For all 108 AI-generated discharge summaries, the total API cost was $7.60 USD.

## Discussion

We found that the AI-generated discharge summaries significantly outperformed actual human written summaries in both quality and readability/conciseness and were comparable to humans in factuality and completeness. This is clinically important because it addresses concerns of inadequate factuality and completeness for use in real-world care scenarios, which has been a concern in previous studies^20^. Moreover, our study was conducted using complete records of clinical notes from real-world inpatient hospital encounters, with comparison to the discharge summaries written by hospital physicians, which has not been done in previous work.

Our findings did not suggest that fabrications and omissions were completely eliminated in the AI-generated summaries, but in cases where they did appear, none of them were judged to result in severe extent of patient harm. Furthermore, manual evaluation of the human-written summaries revealed error rates similar to the AI-generated summaries in multiple subcategories, namely medium likelihood of harm and mild/moderate extent of harm, and low likelihood of harm/no extent of harm. Even within subcategories where AI performed worse than humans, such as mild/moderate extent of harm for omissions, only a small minority of the AI omissions resulted in high likelihood of patient harm. Interestingly, the only omission judged to result in severe patient harm originated from a human-written summary.

For both the human-written and AI-generated discharge summaries, the most common error category was follow-up/recommendations, followed by diagnoses, medications and test results. Williams et al. also found follow-up/recommendations to be one of the most common error categories^23^; our results support that finding. One reason for this order could be that medications and test results are atomic data that are written in a relatively uniform manner, whereas the other two categories can be written in a variety of ways. Since LLMs predict the next word based on inputted information, models may more easily recognize medications and test results, leading to fewer fabrications and omissions in such categories versus follow-up/recommendations and diagnoses. Given the importance of follow up on transition of care and the relative weakness of LLMs to appropriately generate recommendations, it may be important to iterate on prompts and methods to improve LLM-generated follow up instructions.

Quality, readability and conciseness are extremely important dimensions of discharge summaries during the transition of patient care because the receiving provider needs to follow through with the care plan to reduce adverse consequences of an inadequate handoff, such as readmission. Higher readability and quality may reduce the cognitive burden of a receiving provider, so that they can focus on following through with post-hospitalization care and recommendations. LLMs may also shift the burden of writing discharge summaries to the less burdensome task of editing, where discharging providers could simply correct any fabrications or omissions prior to signing LLM-generated discharge summaries. This study has empirically shown that LLMs can write higher-quality and more readable/concise discharge summaries, and can produce outputs in seconds, potentially saving hours of clinicians’ time that could be reallocated to direct patient care.

Much of the previous work on this task, as well as other LLM-based tasks using PHI, has been limited by practical roadblocks such as obtaining full sets of clinical documentation and accessing API-only models in a manner that respects patient privacy laws (i.e., HIPAA), since API usage in this context necessitates sending PHI over the internet to a remote server. Other such roadblocks include filtering and preprocessing clinical documentation at scale, which tends to be very heterogeneous, and cost considerations. This study provides a pilot and proof of concept on preliminary methods for addressing each of these barriers to implementation, bringing us closer to real-life clinical integration of AI-based solutions.

One limitation of this study is that the system prompt used to generate the AI discharge summaries was nonspecific on some of the exact elements that should have been included, such as lists of discharge diagnoses and medications, leading the clinicians performing the manual evaluations to detect omissions that may have been avoided with more specific prompting. Another limitation is that we did not perform data compression (i.e., reducing tokens while preserving information) on the clinical notes, thus limiting the eligible encounters to those with total EHR character lengths under a certain threshold, in this case 120,000. We intend to improve on both issues in future work.

### Future Work

For future work, we will consider iterative approaches to improving quality and reducing errors, particularly for follow-up/recommendations, using few-shot prompting and reflective agentic workflows^25^. Also, given that the previous work on AI discharge summaries in Psychiatry found LLM outputs to be of poor quality, we will use our current methods to explore performance in Psychiatry encounters, which are different from IM encounters in content and style of documentation^22^. Finally, given the human-intensive effort required to manually review and compare discharge summaries, we will explore the use of LLMs to evaluate clinical documentation and detect errors of fabrication and omission to reduce the human effort in evaluation.

## Conclusion

In this pilot study, we found that GPT-4o generated significantly better discharge summaries than human clinicians on dimensions of quality, readability and conciseness, and with similar performance on dimensions of factuality and completeness. Our results suggest that while AI-generated discharge summaries do contain some errors, the error rates are similar to those of humans and the vast majority of them are of minimal clinical consequence. Automating clinical documentation tasks is important because it can save hours of clinicians’ time, reducing burnout and improving care quality. The methods used in this study, including the setup of HIPAA-compliant API access to OpenAI’s latest models and the preprocessing of heterogeneous free-text notes, will inform future iterations of this project and its potential implementation in real world settings.

### Student Paper Disclosure

Author 1 performed all the data processing, analysis and manuscript writing, and is a Computer Science Ph.D. student enrolled at Stony Brook University in Stony Brook, NY, advised primarily by author 9. Authors 2, 3, 4 and 10 conducted the manual evaluations. Authors 5-8 assisted with the clinical aspects of concept work leading to the development of this paper. Authors 9 and 10 provided suggestions and edits for the manuscript.

## Data Availability

All data analyzed in the present study is private health information and is not available for distribution.

